# Incidence of Mortality and Predictors Among Patients with Shock Managed in the Emergency Room of a Large Tertiary Referral Hospital in Ethiopia

**DOI:** 10.1101/2024.02.10.24302628

**Authors:** Kalsidagn Girma Asfaw, Abel Getachew Adugna, Nahom Mesfin Mekonen, Tigist Workneh Leulseged, Merahi Kefyalew Merahi, Segni Kejela, Fekadesilassie Henok Moges

**Affiliations:** Department of Emergency Medicine and Critical Care, Addis Ababa University, College of Health Sciences, Addis Ababa, Ethiopia; Ethio Scandic Internal Medicine Speciality Clinic, Addis Ababa, Ethiopia; Medical Research Lounge, Addis Ababa, Ethiopia; Department of Internal Medicine, St. Paul’s Hospital Millennium Medical College, Addis Ababa, Ethiopia; Department of Surgery in Addis Ababa University College of Health Sciences, Addis Ababa, Ethiopia; Department of Internal Medicine, Aurora West Allis Medical Center, Milwaukee, WI, USA

**Author notes:** **Corresponding author:** Kalsidagn Girma Asfaw.

**Keywords:** Shock, Mortality, Emergency care, Retrospective chart review, Robust poisson regression, Ethiopia

## Abstract

**Background:** Shock is a common emergency condition which can lead to organ failure and death if not diagnosed and managed timely. Despite its huge global impact, data is scarce in resource-limited settings, such as Ethiopia, which hinders the provision of quality care for improved patient outcomes. Hence, the aim of the study was to determine the incidence of death and predictors among adult patients with shock managed at the Emergency Department of St. Paul’s Hospital Millennium Medical College in Ethiopia.

**Methods:** A retrospective chart review study was conducted between July to September 2022 among 178 eligible adult patients who were managed at hospital between October 2021 and May 2022. The characteristics of the participants were summarized using frequency and median with interquartile range. The incidence of mortality was estimated using incidence density using person hour (PH) of observation. To identify predictors of mortality, a generalized linear model using poisson regression model with robust standard errors was run at 5% level of significance, where adjusted relative risk (ARR) with its 95% CI was used to interpret significant results

**Result:** The incidence of death was 6.87 deaths per 1000 PH (95% CI= 5.44 to 8.69). Significant predictors of death were being triaged orange (ARR=0.46, 95% CI=0.24-0.88, p=0.020), having a high shock index (ARR=1.59, 95% CI=1.07-2.36, p=0.021), being diagnosed with septic shock (ARR=3.66, 95% CI=1.20-11.17, p=0.023), taking vasopressors (ARR=3.18, 95% CI=1.09, 9.27, p=0.034), and developing organ failure (ARR=1.79, 95% CI=1.04-3.07, p=0.035).

**Conclusion:** The incidence of mortality among shock patients was found to be considerable but relatively lower than previous studies. To optimize patient care and improve outcomes, it is important to remain vigilant in the proper triage and early diagnosis of shock using more sensitive tools for prompt identification of high-risk cases, as well as to provide timely, prioritized and effective interventions.

## BACKGROUND

Shock is a condition that occurs when the body is unable to deliver enough oxygen to its tissues due to circulatory insufficiency, resulting in an imbalance between tissue oxygen delivery and consumption. Shock can be caused by a variety of factors, including intravascular fluid loss, heart failure, infections, allergies, and physical blockages (1). It is a common medical emergency, accounting for 0.4% to 1.3% of all patients presenting to emergency departments (EDs) (2, 3). The percentage of different types of shock varies depending on the population served by the ED (4). Furthermore, shock, particularly septic shock, is reported to account for up to 9.4% of Intensive Care Unit (ICU) admission (5, 6).

If not diagnosed and managed early, shock is a life-threatening condition with a potential to result in irreversible damage, including multi-organ dysfunction and failure, ultimately leading to death. It is also one of the major causes of mortality in the emergency room (1, 7). Various studies conducted in different clinical settings around the world, including reviews, have shown that the rate of shock-related mortality ranges from 12% to 56.2%, depending on the type of shock (8-13), the severity of shock (14-16), and the severity of underlying illness (15, 17, 18).

Increased risk of death is reported to be associated with severe types of shocks, primarily septic and cardiogenic shocks (9-11, 14, 16), complication with vital organ dysfunction, and delays in presentation (17, 19). In addition, shock index (SI) is reported to be a strong predictor of mortality (20-24). Moreover, the duration of follow-up significantly affects the rate of death, with the risk increasing with follow-up time, peaking at the 7-day and 30-day follow-up periods (8, 24, 25).

Despite the recognized global burden of shock, data on the pattern and outcome of this condition in Ethiopia remain scarce, particularly in the emergency setting where the first point of care is provided. Few studies conducted among ICU patients in Ethiopia have shown that septic shock accounts for 11% of mortality among medical ICU patients (26). Additionally, the mortality rate among patients with shock admitted to the ICU was 51.8%, and the 28-day mortality rate among septic shock patients was 58.3% (27, 28). These findings highlight that shock is also a major problem in the Ethiopian critical care setting and requires further investigation to guide the healthcare system towards providing quality care focused on early recognition, rapid diagnosis, and empiric management of shock patients who presented to the emergency room for improved outcomes and reduced healthcare burden. Hence, the aim of the study was to determine the incidence of death and predictors among adult patients with shock managed at the Emergency Department of St. Paul’s Hospital Millennium Medical College in Ethiopia between October 2021 and May 2022.

## METHODS

### Study Setting and Design

A retrospective chart review study was conducted between July to September 2022 among adult patients with shock who were managed at the adult ED of St. Paul’s Hospital Millennium Medical College (SPHMMC) in Addis Ababa, Ethiopia between October 2021 and May 2022. SPHMMC is one of the largest tertiary referral hospitals in the country with an average annual patient flow of 200,000 patients and a catchment population of 5 million. The adult emergency department is one of the departments which give in-patient medical service for an average of 740 patients monthly. The ED mainly focuses on medical and surgical patients; and trauma patients are seen at another branch of the hospital, Aabet hospital, sited 2.5km away from the main hospital [29].

### Population and Sample Size

All adult patients who visited the ED of the hospital between October 2021 and May 2022 and were diagnosed to have circulatory shock as signed by an Emergency and Critical Care Medicine (ECCM) senior physician or resident were identified to be eligible to be included in the study. Accordingly, a total of 196 patients with shock were identified. From which, 18 patients with incomplete records of major exposures or outcome on their medical charts were excluded. Finally, a total of 178 eligible patients were included in the study. Since the total number of eligible participants identified were small, all of them were included in the study.

### Operational Definitions

#### Shock

Is diagnosed when SBP is less than 90 mmHg or with systemic signs of hypoperfusion with evidence of risk factors to develop shock. It is classified into different types as follows (30).

#### Hypovolemic Shock

Is diagnosed when a patient is in shock as a result of intravascular volume loss due to gastrointestinal, renal, blood or other body fluid loss.

#### Septic Shock

When a patient with evidence or suspected infection remains in shock despite adequate fluid resuscitation.

#### Cardiogenic shock

Is diagnosed when there is decreased cardiac output despite adequate intravascular volume with confirmed underlying cardiac disease.

#### Anaphylactic Shock

patient who has a history of allergy to a known or unknown offender or who for the first time presents with shock after taking a specific allergenic substance whose shock couldn’t be attributed to other causes of shock category.

#### Mixed shock

Is the presence of more than one type of shock.

**Undifferentiated shock** - refers to the situation where shock is recognized but the cause is unclear.

### Data Collection Procedures and Quality Assurance

A pretested data abstraction tool was used to extract data from the medical records of eligible participants. Data on baseline sociodemographic and clinical parameters were collected by one General Practitioner and two ECCM residents after they were trained on the tool with the supervision of a senior ECCM resident. The electronic data was then exported into SPSS software version 25.0 for data management and analysis. Before initiation of analysis, the collected data was managed through data cleaning, transformation, and cross-referencing of inconsistencies.

### Statistical Analysis

The socio-demographic and clinical characteristics of the study participants were summarized using frequency and median with interquartile range (due to the skewed nature of the data as evidenced by Kolmogorov-Smirnov test of normality with a p-value of <0.05). The incidence of mortality was estimated using incidence density with its 95% CI by taking the number of deaths and the total person hour (PH) observation of the study participants.

To identify predictors of mortality, a generalized linear model using poisson regression model with robust standard errors was run. To identify the variables to be fitted in the final model, a univariate analysis was run at a significance level of 25%. The variables that passed the univariate analysis selection criteria and those that were considered to be clinically relevant were then fitted into the final multivariable robust poisson regression model at 5% level of significance. Significant results from the final model were then interpreted using adjusted relative risk (ARR) with its 95% CI. The fitness of the final model was checked using an omnibus test and it showed that the data fitted the model adequately (p-value = 0.006).

## RESULTS

### Baseline Socio-demographic and Clinical Characteristics

Over two-third of the participants were 40 years and older, with the majority being 60 years and older (39.9%). Ninety-one (51.1%) were females and 78 (43.8%) were residents of Addis Ababa. The majority (63.0%) were referred from another institution, mainly a health center (32.0%), followed by self-referral for those who already have follow-up at the hospital (25.8%), and the rest 20 (11.2%) were patients who were linked from the outpatient department (OPD) of the hospital itself. For the referred cases, the median time from referral to arrival was 4 hours (IQR, 2-9.25 hours).

Upon presentation to the ER, the majority 101 (56.7%) were triaged to be red, followed by yellow (21.9%), orange (14.0%), and green (7.3%). One hundred twenty-two (68.5%) of the patients claimed to have symptoms less than one week. Over two-third (70.8%) had one or more comorbid illness of which malignancy accounted for the majority documented in 31 (24.6%) cases, followed by chronic liver disease in 23 (18.3%), and HIV in 13 (10.3%). Nine (7.1%) patients had multiple comorbidities.

The majority of patients presented with a deranged vital sign. Most patients had low blood pressure with systolic blood pressure (SBP) below 90 mmHg for 149 patients (83.7%), and 6 diastolic blood pressure (DBP) between 40 and 59 mmHg for 99 patients (55.6%). Pulse rate (PR) was elevated in over half, with 101-120 beats per minute (bpm) in 54 (30.3%), and >120 in 41 (23.0%). Respiratory rate (RR) was below 24 breaths per minute for most (66.9%). Shock index (SI) was elevated in a significant proportion, 134 patients (75.3%) had an SI of 1 or higher, while only 10 patients (5.6%) had an SI below 0.7. **(Table 1)**

**Table 1:** Baseline socio-demographic and clinical characteristics among adult patients with shock managed at ED of SPHMMC in Ethiopia, October 2021 to May 2022 (n=178)

| Variable | Frequency (%) | Variable | Frequency (%) |
| --- | --- | --- | --- |
| **Age category** |  | **Comorbid illness** |  |
| <40 | 54 (30.3) | No | 52 (29.2) |
| 40-59 | 53 (29.8) | Yes | 126 (70.8) |
| ≥ 60 | 71 (39.9) | **SBP** |  |
| **Sex** |  | <90 | 149 (83.7) |
| Male | 87 (48.9) | ≥ 90 | 29 (16.3) |
| Female | 91 (51.1) | **DBP** |  |
| **Place of residence** |  | <40 | 38 (21.3) |
| Addis Ababa | 78 (43.8) | 40-59 | 99 (55.6) |
| Outside Addis Ababa | 100 (56.2) | 60-90 | 41 (23.0) |
| **Referred from** |  | **MAP** |  |
| Health center | 57 (32.0) | <65 | 140 (78.7) |
| Primary hospital | 19 (10.7) | ≥ 65 | 38 (21.3) |
| General hospital | 29 (16.3) | **PR** |  |
| Private center | 7 (3.9) | <60 | 16 (9.0) |
| Self-referral | 46 (25.8) | 60-100 | 67 (37.6) |
| Linked from OPD | 20 (11.2) | 101-120 | 54 (30.3) |
| **Initial triage category** |  | >120 | 41 (23.0) |
| Green | 13 (7.3) | **RR** |  |
| Yellow | 39 (21.9) | ≤ 24 | 119 (66.9) |
| Orange | 25 (14.0) | 25 - 34 | 42 (23.6) |
| Red | 101 (56.7) | ≥ 35 | 17 (9.6) |
| **Symptom duration** |  | **SI** |  |
| < 7 days | 122 (68.5) | ≤ 0.7 | 10 (5.6) |
| ≥ 7 days | 56 (31.5) | 0.7 - 1.0 | 34 (19.1) |
|  |  | ≥ 1 | 134 (75.3) |

### Laboratory and POCUS findings

The majority of the patients had a deranged complete blood count result, including mild to moderate anemia in 35 (19.7%), severe anemia in 66 (37.1%), leukopenia in 23 (12.9%), leukocytosis in 76 (42.7%), mild to moderate thrombocytopenia in 25 (14.0%), and severe thrombocytopenia in 104 (58.4%). Electrolyte derangement was also reported in over a third of the patients, mainly hyponatremia in 60 (33.7%), followed by hypokalemia in 53 (29.8%), hyperkalemia in 17 (9.6%), and hypernatremia in 13 (7.3%). Close to half (44.9%) of the patients had a raised creatinine level above 1.2.

Point-of-Care Ultrasound (POCUS) was done for 154 patients and revealed that the majority of the patients had a collapsed or kissing IVC (73.4%) and good cardiac contractility (70.8%). Nearly 30% of patients had some type of fluid collection, mainly pleural collection in 20 (12.9%) and ascites in 20 (12.9%). Multiple B-lines and consolidation was observed in only 49 (29.9%) patients. **(Table 2)**

**Table 2:** Laboratory and POCUS findings of adult patients with shock managed at ED of SPHMMC in Ethiopia, October 2021 to May 2022 (n=178)

| Variable |  | Frequency (%) | Variable | Frequency (%) |
| --- | --- | --- | --- | --- |
| Laboratory parameters (n=178) |  |  | POCUS finding (n=154) |  |
| **Hg b** | Normal | 77 (43.2) | **IVC status** |  |
|  | Mild to moderate anemia | 35 (19.7) | Collapsed/kissing | 113 (73.4) |
|  | Severe anemia | 66 (37.1) | Full | 27 (17.5) |
| **W BC** | Normal | 79 (44.4) | Dilated/plethoric | 14 (9.1) |
|  | Leucopenia | 23 (12.9) | **Cardiac contractility status** |  |
|  | Leukocytosis | 76 (42.7) | Good | 109 (70.8) |
| **PL T** | Normal | 49 (27.5) | Poor/weak | 39 (25.3) |
|  | Mild to moderate thrombocytopenia | 25 (14.0) | Hyperdynamic | 6 (3.9) |
|  | Severe Thrombocytopenia | 104 (58.4) | **Fluid collection** |  |
| **Na** | Normal | 105 (59.0) | No collection | 107 (69.5) |
|  | Hyponatremia | 60 (33.7) | Pleural collection | 20 (12.9) |
|  | Hypernatremia | 13 (7.3) | Ascites | 20 (12.9) |
| **K** | Normal | 108 (60.7) | Pericardial collection | 2 (0.01) |
|  | Hypokalemia | 53 (29.8) | Pleural and peritoneal collection | 3 (0.02) |
|  | Hyperkalemia | 17 (9.6) | Pleural and pericardial collection | 2 (0.01) |
| **Cr** | <0.6 | 35 (19.7) | **Multiple B-lines and Consolidation** |  |
|  | 0.6-1.2 | 63 (35.4) | No | 108 (70.1) |
|  | >1.2 | 80 (44.9) | Yes | 46 (29.9) |

### Type of Shock and Management

The commonly diagnosed types of shocks were hypovolemic shock in 75 (42.1%) and septic shock in 73 (41.0%) patients. This was followed by cardiogenic shock which was diagnosed in 13 (7.3%). Another 13 patients (9.6%) presented with a mixed type of shock including septic and hypovolemic in 8 (4.5%), and septic and cardiogenic in 5 (2.8%). Moreover, 3 patients (1.7%) were diagnosed with obstructive shock, and 1 patient (0.6%) had anaphylactic shock. Shock was diagnosed at presentation for 145 patients (81.5%) and the rest 33 patients (18.5%) were diagnosed after admission after a median of 18 hours (minimum of 2 hours and maximum of 96 hours). From the 33 patients, the majority were those who were triaged as yellow (14/33) or green (9/33) on admission.

Upon admission, 159 (89.3%) of patients required fluid resuscitation and 43 (24.2%) needed blood transfusion. Antibiotics were administered for 138 (77.5%) patients. Additionally, vasopressors and steroids were administered for 98 (55.1%) and 66 (37.1%) patients, respectively.

Close to half of the patients (48.9%) developed organ failure, mainly renal failure which was documented in 40 (46.0%) patients. In addition, 36 (41.4%) patients had multiple organ failure (MOF). Furthermore, 52 (29.2%) patients had electrolyte imbalance, mainly hypokalemia (52.9%), followed by hyponatremia (29.4%), and hyperkalemia (13.7%). **(Table 3)**

**Table 3:**
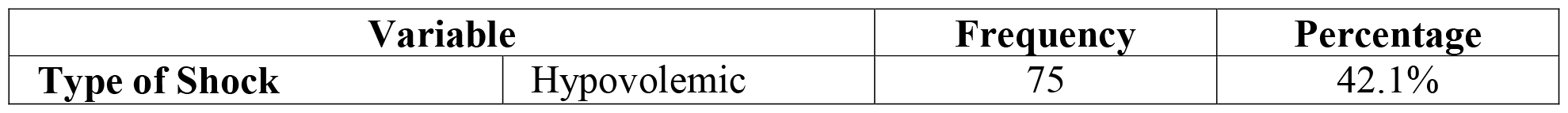
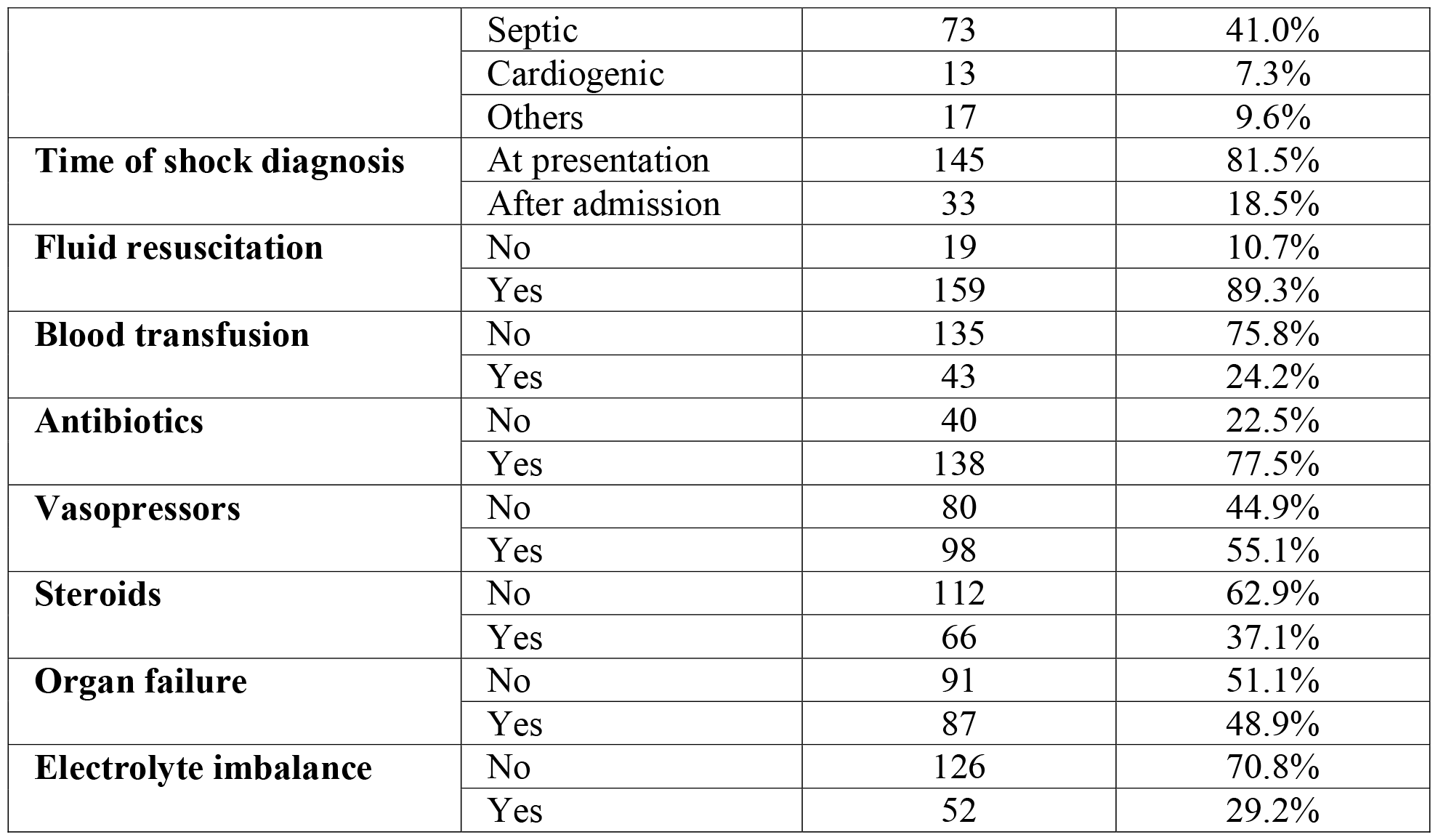
Type of shock and management provided among adult patients with shock managed at ED of SPHMMC in Ethiopia, October 2021 to May 2022 (n=178)

### Incidence of Mortality

The study participants were observed for a median of 48 hours (IQR, 24-72 hours). During this time, 70 of the 178 hospitalized patients died, resulting in an incidence density of 6.87 deaths per 1000 PH (95% CI= 5.44 to 8.69). The major causes of death were refractory septic shock in 35 (19.7%) patients and MOF in 21 (11.8%) patients. During the course of treatment, 28 patients needed further in patient follow-up, hence 20 (11.2%) were admitted to the ward and 8 (4.5%) were admitted to the ICU.

### Predictors of Mortality

A multivariable robust poisson regression analysis was run by fitting the following exposure variables; age group, sex, triage category, comorbid illness, SBP, SI, Hgb, WBC, PLT, Na, K, Cr, type of shock, time of shock diagnosis, IV fluid, blood transfusion, antibiotics, vasopressors, and organ failure. Subsequently, triage category, SI, type of shock, vasopressor use, and organ failure were found to be significant predictors of mortality in shock patients.

The risk of death was found to be lower by 54% among patients who were triaged orange (**ARR=0.46, 95% CI=0.24-0.88, p=0.020**) as compared to patients who were triaged green on admission. Patients with SI of 1 or higher were 59% more likely to die than those with SI of less than 1 (**ARR=1.59, 95% CI=1.07-2.36, p=0.021**).

The risk of death among patients who were diagnosed with septic shock and those who required vasopressors was 3.66 times (**ARR=3.66, 95% CI=1.20-11.17, p=0.023**) and 3.18 times (**ARR=3.18, 95% CI=1.09, 9.27, p=0.034**) higher than those who presented with hypovolemic shock and those who did not require vasopressor treatment, respectively. Furthermore, patients who developed organ failure had a 79% increased risk of death than those who did not develop such complications (**ARR=1.79, 95% CI=1.04-3.07, p=0.035**). **(Table 4)**

**Table 4:**
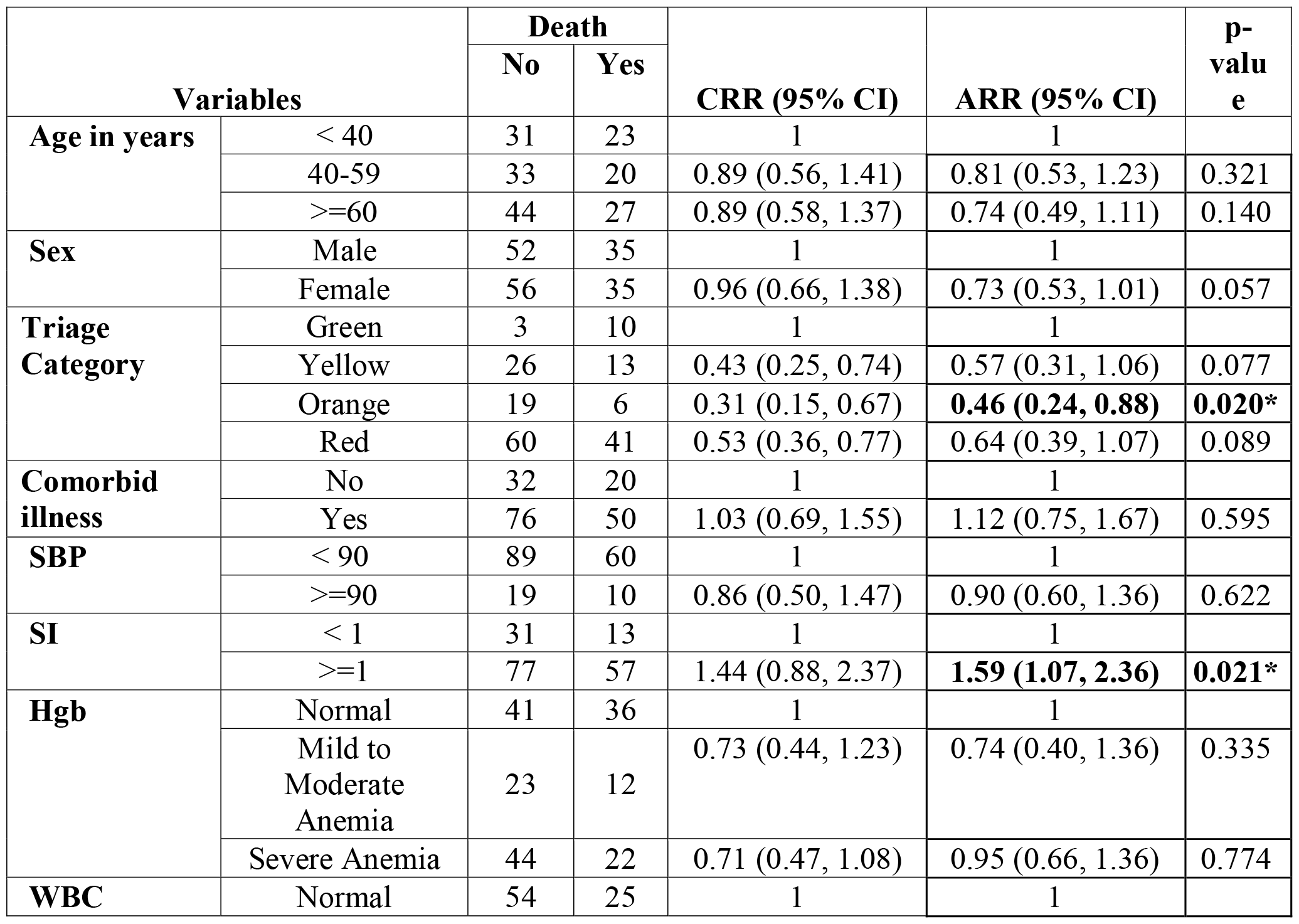
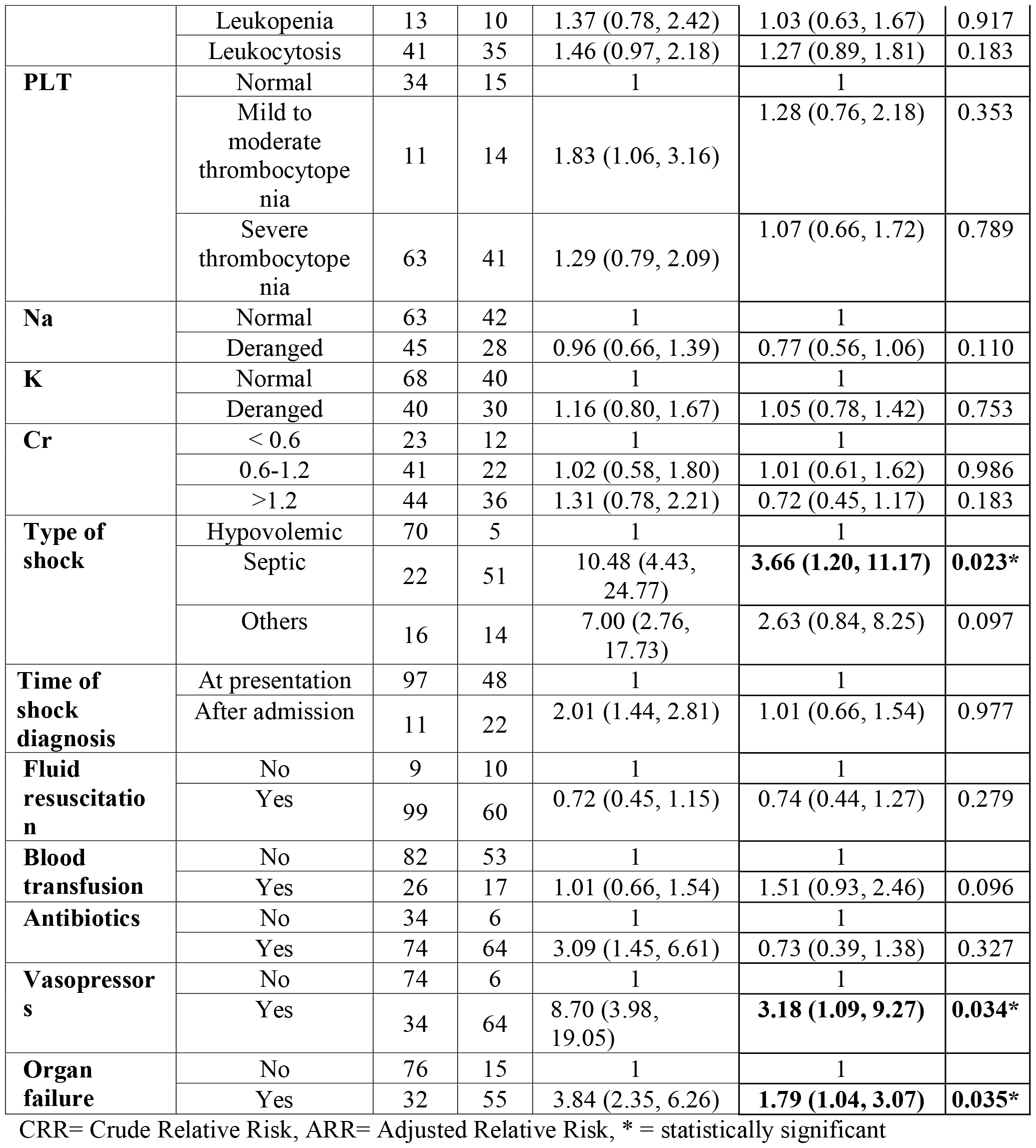
Predictors of mortality among adult patients with shock managed at ED of SPHMMC in Ethiopia, October 2021 to May 2022 (n=178)

## DISCUSSION

This study investigated the incidence and predictors of death among 178 adult patients with shock managed at the ED of a large tertiary referral hospital in Ethiopia between October 2021 and May 2022. The incidence of morality was 6.87 deaths per 1000 PH (95% CI = 5.44 to 8.69), which is a significant death rate, but it is found to be relatively lower than previous reports. A death rate of 12% to 32% was reported in studies conducted in similar settings and developed health care settings, with the risk increasing by up to 50% for patients who presented with severe underlying medical conditions and those who developed complications during the course of treatment (2, 26). This decreased rate could be due to the relatively lower proportion of patients who were triaged to be red as compared to the other studies.

Significant predictors of mortality were found to be triage category, SI, type of shock, use of vasopressors, and complication with organ failure. Accordingly, the risk of death was found to be lower by 54% among patients who were triaged orange as compared to patients who were triaged green on admission. This could be due to the fact that within the emergency room setting, where resources and attention may be disproportionately allocated to patients with perceived higher acuity, especially in a resource limited setting, those triaged less urgent may face challenges in accessing timely evaluation and treatment, thus potentially compromising their outcomes. Furthermore, patients who were triaged orange represent patients with conditions serious enough to warrant prompt attention but not immediately life-threatening as those triaged red. Hence, early evaluation and intervention for their conditions could contribute to improved outcomes and lower mortality (31). Moreover, timing of shock diagnosis is critical, as delayed diagnosis is likely associated with poor outcome. The study found that 33 (18.5%) patients were diagnosed with shock after admission, within a median of 18 hours, from which the majority were triaged yellow (14/33) or green (9/33) at arrival. This shows that higher mortality rates in these groups could be directly related to late shock diagnosis and subsequently delayed care.

SI of 1 or higher was associated with a 59% increased risk of death than those with SI of less than 1. High SI indicates both low blood pressure and high heart rate, exacerbating the detrimental effects on tissue oxygenation and organ function, and thereby increasing the risk of death. The finding aligns with previous research reports across various clinical scenarios demonstrating that elevated SI is associated with increased morbidity and mortality. Moreover, studies revealed that SI also has superiority over traditional parameters such as SBP alone as a crucial prognostic marker through improving the identification of patients requiring urgent care, optimizing resource allocation, and potentially reducing under-triage rates without significantly increasing over-triage (20-24).

Presenting with septic shock was associated with an almost four-fold increased risk of death than presenting with hypovolemic shock. Septic shock is caused by a systemic inflammatory response to infection and is characterized by a complex cascade of inflammatory mediators, ultimately leading to failure in multiple organs. As a result, even with prompt medical intervention, patients with septic shock face a significantly higher risk of complications and death. This increased mortality rate in septic shock is also well established in other studies with varying levels of risk (27, 28, 32-35).

Use of vasopressor was associated with a three-fold increased risk of death. The need for vasopressor therapy indicates a significant hemodynamic instability and a challenging clinical situation. It also often reflects the presence of underlying medical conditions, advanced illness, or delayed medical attention, all of which are associated with worse outcomes. This association between vasopressor use and increased mortality has been consistently observed in various clinical settings and across different studies as well (36, 37).

Finally, developing organ failure was associated with a 79% increased risk of death. This could be because organ failure can alter critical metabolic processes and impair the immune system, resulting in cellular damage and increased susceptibility to infection, ultimately increasing the risk of mortality. This risk increases further with the severity of the initial insult, the presence of pre-existing comorbidities, and delays in receiving proper care. This finding is also consistent with previous studies (14-16, 17, 19).

The study adds to the current understanding in the field, considering scarcity of evidence in similar settings, and will serve as a base for future research. However, the following limitations necessitate cautious interpretation of the findings. Due to the retrospective nature of the study, additional relevant variables were not controlled in the study including details on patient management, such as specific medication dosages and the adequacy of overall care, including proper fluid resuscitation. Additionally, information on underlying medical conditions, such as medication usage, adherence, and control levels, as well as behavioral factors, were not included. Furthermore, while the study was conducted in a large referral hospital with a high patient volume, its single-center design limits the generalizability of the findings to similar settings.

## CONCLUSION

The incidence of mortality among shock patients was found to be considerable but relatively lower than previous studies. Significant predictors of death were being triaged green, having a high shock index, being diagnosed with septic shock, taking vasopressors, and developing organ failure. Therefore, it is important to remain vigilant in the proper triage and early diagnosis of shock using more sensitive tools for prompt identification of high-risk cases, as well as to provide timely, prioritized and effective interventions to optimize patient care and improve outcomes. For more generalizable results applicable to a wider clinical setting, a multicenter prospective study investigating all relevant patient and clinical characteristics is recommended.

## Data Availability

All relevant data are available upon reasonable request from Kalsidagn Girma Asfaw at.

## Declaration

### Ethics approval and consent to participate

The study was carried out after obtaining ethical approval from the institutional review board of St. Paul’s Hospital Millennium Medical College (SPHMMC-IRB). The St. Paul’s Hospital Millennium Medical College institutional review board (SPHMMC-IRB) also waived the need for informed consent since the study used secondary data (Ref. No. PM23/62). The study was carried out in accordance with relevant guidelines and regulations. Confidentiality of the patient’s information contained within their registration books, cards, and referral papers were assured by using of medical record number and omitting any personal identifiers. Furthermore, access to the collected information was limited to the investigators.

## Consent for publication

Not applicable

## Availability of data and materials

All relevant data are available upon reasonable request from Kalsidagn Girma Asfaw at.

## Authors’ Contribution

KGA, AGA, and NMM conceived and designed the study. TWL, MKM, SK, and FHM contributed to the conception and design of the study. KGA and TWL performed statistical analysis, and drafted the initial manuscript. AGA and NMM contributed to the interpretation of the findings and drafting of the manuscript. MKM, SK, and FHM revised the manuscript. All authors approved the final version of the manuscript.

## Acknowledgment

The authors would like to thank all individuals involved in the data collection, supervision, and facilitation of the research work.

## REFERENCES

1. Russell JA, Rush B, Boyd J. Pathophysiology of Septic Shock. Critical care clinics. 2018;34(1):43–61.

2. Holler JG, Jensen HK, Henriksen DP, et al: Etiology of shock in the emergency department; a 12 year population based cohort study. Shock. 51:60, 2019. [PMID: 27984523]

3. Holler JG, Bech CN, Henriksen DP, Mikkelsen S, Pedersen C, Lassen AT (2015) Nontraumatic Hypotension and Shock in the Emergency Department and the Prehospital setting, Prevalence, Etiology, and Mortality: A Systematic Review. PLoS ONE 10(3): e0119331.

4. Kheng CP, Rahman NH. The use of end-tidal carbon dioxide monitoring in patients with hypotension in the emergency department. International journal of emergency medicine. 2012;5(1):31.

5. Mbengono JAM, Tochie JN, Ntock FN, Bertrand Nzouango Y, Kona S, Ateba GN, et al. The Epidemiology, Therapeutic Patterns, Outcome, and Challenges in Managing Septic Shock in a Sub-Saharan African Intensive Care Unit: A CrossSectional Study. Hospital Practices and Research. 2019;4(4):117–21.

6. Artero A, Zaragoza R, Camarena JJ, Sancho S, Gonzalez R, Nogueira JM. Prognostic factors of mortality in patients with community-acquired bloodstream infection with severe sepsis and septic shock. Journal of critical care. 2010;25(2):276–81.

7. David F Gaieski MD, Mark E Mikkelsen MD, MSCE. [Internet]. UpToDate. [cited 2022Apr10]. Available from: https://www.uptodate.com/contents/definition-classification-etiology-and-pathophysiology-of-shock-in-adults?search=hypovolemicshock&source=search_result&selectedTitle=2~150&usage_type=default&display_rank=2

8. Gitz Holler J, Jensen HK, Henriksen DP, et al. Etiology of Shock in the Emergency Department: A 12-Year Population-Based Cohort Study. Shock. 2019;51(1):60–67.

9. Guly HR, Bouamra O, Lecky FE, Trauma A, Research N. The incidence of neurogenic shock in patients with isolated spinal cord injury in the emergency department. Resuscitation. 2008;76(1):57–62.

10. Fleischmann C, Scherag A, Adhikari JKJ, et al: Assessment of global incidence and mortality of hospital-treated sepsis. Current estimates and limitations. Am J Resp Crit Care Med 193: 259, 2016. [PMID: 26414292]

11. Jeger RV, Radovanovic D, Hunziker PR, et al: Ten-year trends in the incidence of cardiogenic shock. Ann Intern Med 149: 618, 2008. [PMID: 18981487]

12. Joakyna De Santiago MR-F. Early Diagnosis of Pediatric Laryngotracheal Rupture Following Minor Blunt Trauma. Tropical Medicine & Surgery. 2013;01(05).

13. Becker, J. U., Theodosis, C., Jacob, S. T., Wira, C. R., & Groce, N. E. (2009). Surviving sepsis in low-income and middle-income countries: new directions for care and research. The Lancet Infectious Diseases, 9(9), 577–582. doi:10.1016/s1473-3099(09)70135-5

14. Su L, Ma X, Rui X, He H, Wang Y, Shan G, et al. Shock in China 2018 (SIC-study): a cross-sectional survey. Annals of translational medicine. 2021;9(15):1219.

15. Artero A, Zaragoza R, Camarena JJ, Sancho S, Gonzalez R, Nogueira JM. Prognostic factors of mortality in patients with community-acquired bloodstream infection with severe sepsis and septic shock. Journal of critical care. 2010;25(2):276–81.

16. Mbengono JAM, Tochie JN, Ntock FN, Bertrand Nzouango Y, Kona S, Ateba GN, et al. The Epidemiology, Therapeutic Patterns, Outcome, and Challenges in Managing Septic Shock in a Sub-Saharan African Intensive Care Unit: A CrossSectional Study. Hospital Practices and Research. 2019;4(4):117–21.

17. Holler JG, Bech CN, Henriksen DP, Mikkelsen S, Pedersen C, Lassen AT (2015) Nontraumatic Hypotension and Shock in the Emergency Department and the Prehospital setting, Prevalence, Etiology, and Mortality: A Systematic Review. PLoS ONE 10(3): e0119331.

18. Mbengono JAM, Tochie JN, Ntock FN, Bertrand Nzouango Y, Kona S, Ateba GN, et al. The Epidemiology, Therapeutic Patterns, Outcome, and Challenges in Managing Septic Shock in a Sub-Saharan African Intensive Care Unit: A CrossSectional Study. Hospital Practices and Research. 2019;4(4):117–21.

19. Tintinalli, J. E. (2019). chapter 12. In Tintinalli’s emergency medicine: A comprehensive study guide, 9th edition. essay, McGraw-Hill Education.

20. Liu, Y. (2012). Modified shock index and mortality rate of emergency patients. World Journal of Emergency Medicine, 3(2), 114. doi:10.5847/wjem.j.issn.1920-8642.2012.02.006

21. Koch E, Lovett S, Nghiem T, Riggs RA, Rech MA. Shock index in the emergency department: utility and limitations. Open Access Emerg Med. 2019 Aug 14;11:179–199. doi: 10.2147/OAEM.S178358. PMID: 31616192; PMCID: PMC6698590.

22. Middleton DJ, Smith TO, Bedford R, Neilly M, Myint PK. Shock Index Predicts Outcome in Patients with Suspected Sepsis or Community-Acquired Pneumonia: A Systematic Review. J Clin Med. 2019 Jul 31;8(8):1144. doi: 10.3390/jcm8081144. PMID: 31370356; PMCID: PMC6723191.

23. El-Menyar A, Al Habib KF, Zubaid M, Alsheikh-Ali AA, Sulaiman K, Almahmeed W, Amin H, Al Motarreb A, Ullah A, Suwaidi JA. Utility of shock index in 24,636 patients presenting with acute coronary syndrome. Eur Heart J Acute Cardiovasc Care. 2020 Sep;9(6):546–556. doi: 10.1177/2048872619886307. Epub 2019 Nov 8. PMID: 31702396.

24. Bauer M, Gerlach H, Vogelmann T, Preissing F, Stiefel J, Adam D. Mortality in sepsis and septic shock in Europe, North America and Australia between 2009 and 2019-results from a systematic review and meta-analysis. Critical care. 2020;24(1):239.

25. Gupta N, Aggarwal S, Gaglianello N, Bangalore S, Cinquegrani MP. Abstract 13061: Trends in Incidence, Management and Outcomes of Heart Failure Patients with Cardiogenic Shock. Circulation. 2014;130(suppl_2):A13061–A.

26. PATTERN AND OUTCOME OF MEDICAL INTENSIVE CARE UNIT ADMISSIONS TO AYDER COMPREHENSIVE SPECIALIZED HOSPITAL IN TIGRAY, ETHIOPIA | Ethiopian Medical Journal;2018,English,Kibreab Gidey

27. Worku A, Sultan A, Selassie L, Shumet A, G/Selassie K, Bekele A, et al. Shock in an Ethiopian Medical Intensive Care Unit: Characteristics and Outcomes at Tikur Anbessa Specialized Hospital (TASH) in Addis Ababa, Ethiopia. B47 CRITICAL CARE: CLINICAL RESEARCH DISCOVERIES, EPIDEMIOLOGY, AND HEALTH SERVICES FOR SEPSIS. p. A3692–A.

28. Assessment of Incidence and Treatment Outcome of Septic Shock among Patients Admitted to Adult Intensive Care Unit of Tikur Anbessa Specialized Hospital, Addis Ababa, Ethiopia - Africa Thesis Bank

29. En.wikipedia.org. 2022. St. Paul’s Hospital Millennium Medical College - Wikipedia. [online] Available at: <https://en.wikipedia.org/wiki/St._Paul%27s_Hospital_Millennium_Medical_College#cite_note-2> [Accessed 24 March 2022].

30. UpToDate.(n.d.).UpToDate.https://www.uptodate.com/contents/definition-classification-etiology-and-pathophysiology-of-shock-in-adults

31. Johansson, A., Ekwall, A., Forberg, J.L. et al. Development of outcomes for evaluating emergency care triage: a Delphi approach. Scand J Trauma Resusc Emerg Med 31, 10 (2023).

32. Khalil, O., Sakr, M., Selim, F., Mohammed Salem I. PREVALENCE, PATTERN and CLINICAL OUTCOME OF CIRCULATORY SHOCK IN CRITICALLY ILL PATIENTS IN MEDICAL ICU. Zagazig University Medical Journal, 2018; 24(4): 329–337. doi: 10.21608/zumj.2018.13226

33. Metogo Mbengono, J. A., Tochie, J. N., Ndom Ntock, F., Nzoaungo, Y. B., Kona, S., Ngono Ateba, G., Tocko, C., Colibaly, A., Beyiha, G., Ze Minkande, J. The Epidemiology, Therapeutic Patterns, Outcome, and Challenges in Managing Septic Shock in a Sub-Saharan African Intensive Care Unit: A Cross-Sectional Study. Hospital Practices and Research, 2019; 4(4): 117–121. doi: 10.15171/hpr.2019.24

34. Shankar-Hari M, Phillips GS, Levy ML, Seymour CW, Liu VX, Deutschman CS, et al. Developing a New Definition and Assessing New Clinical Criteria for Septic Shock: For the Third International Consensus Definitions for Sepsis and Septic Shock (Sepsis-3). Jama. 2016;315(8):775–87.

35. Singer M, Deutschman CS, Seymour CW, Shankar-Hari M, Annane D, Bauer M, et al. The Third International Consensus Definitions for Sepsis and Septic Shock (Sepsis-3). Jama. 2016;315(8):801–10.

36. Mark, D.G., Morehouse, J.W., Hung, YY. et al. In-hospital mortality following treatment with red blood cell transfusion or inotropic therapy during early goal-directed therapy for septic shock: a retrospective propensity-adjusted analysis. Crit Care 18, 496 (2014).

37. Awad, W.B., Nazer, L., Elfarr, S. et al. A 12-year study evaluating the outcomes and predictors of mortality in critically ill cancer patients admitted with septic shock. BMC Cancer 21, 709 (2021)

